# Effects of maximal speed locomotor training on spatiotemporal gait changes in individuals with chronic stroke: A secondary analysis of a randomized controlled trial

**DOI:** 10.1101/2024.08.27.24312508

**Authors:** Daria Pressler, Sarah M. Schwab-Farrell, Darcy S. Reisman, Sandra A. Billinger, Pierce Boyne

## Abstract

**Objective:** To investigate longitudinal changes in spatiotemporal gait parameters after maximal versus moderate speed locomotor training in chronic stroke, by comparing short-burst high-intensity interval training (HIIT) versus moderate-intensity aerobic training (MAT). Compared to MAT, short-burst HIIT was hypothesized to exhibit greater improvement in non-paretic step length.

**Design:** Secondary analysis from the HIT-Stroke randomized controlled trial

**Setting:** Three rehabilitation research centers

**Participants:** Individuals with chronic stroke and residual walking limitations (N=55)

**Interventions:** Participants were randomized to short-burst HIIT (N=27) or MAT (N=28) for 45 minutes of walking practice, 3 times weekly, over 12 weeks. HIIT involved 30-second bursts of maximum walking speed, targeting >60% heart rate reserve (HRR). MAT involved continuous walking, targeting 40%-60% HRR.

**Main Outcome Measure(s):** Mean spatiotemporal gait parameter changes between groups, averaging the 4-week, 8-week, and 12-week estimates minus baseline. The primary measure of interest was non-paretic step length, an indicator of paretic propulsion and biomechanical efficiency.

**Results:** Non-paretic step length increased significantly more in the HIIT group (+4.4 cm [95% CI, 1.9, 6.9]) compared to the MAT group (+0.1 [-2.5, 2.7]; HIIT vs. MAT p = .01). Both groups demonstrated significant increases in cadence, paretic step length, and bilateral single support time, and significant decreases in the coefficient of variation (CV) for stride velocity, stride time, and stride length. Symmetry measures did not significantly change in either group.

**Conclusions:** Greater increases in non-paretic step length with short-burst HIIT suggest that maximal speed training may yield greater increases in paretic propulsion, a marker of biomechanical efficiency. Both moderate and maximal speed training (MAT and HIIT) appear to reduce spatiotemporal variability, possibly indicating improved gait stability.

## INTRODUCTION

Moderate to high-intensity gait training (M-HIT) involves walking at faster than comfortable speeds to elicit aerobic training intensities (≥40% heart rate reserve). This approach is strongly recommended for enhancing walking capacity (i.e. speed and endurance) in individuals with chronic stroke,^1,2^ and evidence suggests that higher training intensities elicit greater gains.^3^ Walking capacity improvements with M-HIT are partly attributed to adaptive biomechanical changes, notably the restoration of propulsive forces generated by the lower limbs during the terminal stance phase of the gait cycle.^4-7^ These propulsive forces may increase bilaterally after M-HIT, and increased propulsion contribution from the paretic limb is thought to be a key indicator of improved biomechanical efficiency.^8,9^ Paretic propulsion also contributes to non-paretic step length, establishing non-paretic step length as a proxy measure for paretic propulsion.^6,10,11^

Propulsion and biomechanical efficiency have been shown to be greater while individuals with stroke are walking at faster-than-comfortable speeds, and faster training speed during M-HIT appears to be a key mediator of these adaptive biomechanical changes and walking capacity gains.^7,12-17^ However, it remains to be explored whether training at *maximal* speeds could lead to even more pronounced biomechanical improvements. This is because previous studies assessing biomechanical changes from M-HIT have used longer bouts of continuous walking, whereas individuals with stroke can achieve significantly faster speeds during short walking bursts.^18^

Locomotor high-intensity interval training (HIIT) is a method of M-HIT that maximizes training speed using short bursts of walking activity interspersed with rest breaks.^3,18,19^ Despite its potential, no controlled studies have tested changes in paretic limb biomechanics associated with short-burst HIIT. Spatiotemporal gait measures, such as non-paretic step length, may provide valuable preliminary insights into these biomechanical changes, as paretic propulsion influences non-paretic step length, and improvements in paretic propulsion are thought to be an energetically efficient way to increase walking speed, thereby enhancing biomechanical efficiency.^6,10,11^ However, no previous studies have assessed changes in non-paretic step length or other limb-specific spatiotemporal changes following *maximal* speed locomotor training (short-burst HIIT) vs. moderate-intensity aerobic training (MAT).

Therefore, the purpose of this study was to assess changes in non-paretic step length after short-burst HIIT versus MAT in chronic stroke. Compared to MAT, we hypothesized HIIT would exhibit greater increases in non-paretic step length. Additionally, we evaluated the impact of HIIT and MAT on other spatiotemporal measures to provide a more comprehensive understanding of gait changes with these two types of training.

## METHODS

This study analyzed data from the HIT-Stroke Trial, which randomized 55 participants to either short-burst HIIT (N=27) or MAT (N=28).^3^ Target training volume was 45 minutes, 3 times weekly, over 12 weeks, divided into three blocks, each lasting 4 weeks (12-sessions), with outcome testing after each block. The primary HIT-Stroke Trial analysis found a significantly larger improvement in 6-minute walk distance and gait speed for the HIIT group compared to the MAT group.^3^ The current study expanded on these previous findings by analyzing spatiotemporal and gait variability data from the HIT-Stroke Trial. This research was approved by the University of Cincinnati Institutional Review Board, and we used the CONSORT checklist when writing our report.^20^

### Participants

Participants were recruited from the community and provided written informed consent. Inclusion criteria were: age 40-80, single stroke between 6 months and 5 years prior, walking speed of ≥ 1.0 m/s, ability to walk 10 meters over ground without continuous assistance, ability to walk at least 3 minutes on a treadmill at ≥ 0.13m/s (0.3mph), stable cardiovascular condition (American Heart Association class B), and ability to follow instructions and communicate with investigators. Exclusion criteria were: cardiovascular contraindications to vigorous exercise, implanted pacemaker or defibrillator, significant ataxia or neglect (National Institutes of Health Stroke Scale item scores >1), severe lower limb spasticity (Ashworth scale scores >2), foot drop or lower limb instability without adequate stabilization (e.g., ankle foot orthosis), other significant neurological disorders besides stroke, recent history of substance misuse, significant mental illness, unmanaged major poststroke depression (Patient health Questionnaire score ≥10), active participation in other physical therapy or research studies, botulinum toxin injection to the affected lower limb in the past 3 months or planned within the next 4 months, inability to walk outside the home prior to the stroke, other significant medical conditions which could hinder improvement or jeopardize safety, pregnancy, and experience with fast treadmill walking in the past year.

### Intervention & Data Collection

The HIIT group followed a short-interval protocol with 30-second bursts of maximum walking speed, alternated with 30-to 60-second passive rests (standing or seated), targeting above 60% heart rate reserve (HRR). The MAT group performed continuous walking, adjusting speed to maintain 40% ± 5% HRR and incrementally increasing by 5% every two weeks up to 60% HRR as tolerated.

### Spatiotemporal Gait Outcome Testing

Outcomes were assessed at baseline (before randomization) and after 4, 8, and 12 weeks of training. During tests, participants used their usual orthotic and assistive devices. Participants made two passes across a sensor-embedded electronic walkway at a comfortable speed (GaitRite MAT, CIR Systems, Franklin, NJ; or Zeno Walkway, ProtoKinetics LLC, Havertown, PA). Passes were combined, and parameters were averaged across gait cycles. The primary measure of interest was non-paretic step length as a marker of paretic propulsion and indicator of biomechanical efficiency. To provide a more comprehensive view of the impact of HIIT and MAT on spatiotemporal measures, we also assessed changes in variability measures—including standard deviation (SD: absolute variability) and coefficient of variation (CV: relative variability) for stride velocity, time, and length. Additionally, we evaluated outcome data for cadence, single-limb support times, and symmetry measures. The paretic step ratio (relative step length) was calculated using the formula: Paretic / (Paretic + Nonparetic) step length.^6^ Values could range from 0 to 100%, where 50% represents perfect symmetry. Values above 50% indicate a longer non-paretic step length, while values below 50% indicate a longer paretic step length. Absolute symmetry measures were calculated for step length, step time and single support time using the formula: (1 − | Paretic – Nonparetic | / (Paretic + Nonparetic)) ∗ 100%.^21^ Values could range from 0 to 100%, where 0% indicates complete asymmetry and 100% represents perfect symmetry.

### Data Analysis

Linear models were obtained with each spatiotemporal gait parameter as the dependent variable in a separate model. Each model included fixed effects for treatment group, testing time point (baseline, 4-week, 8-week, and 12-week), group-by-time interaction, study site, study site–by-time interaction, baseline walking limitation severity, and baseline walking limitation severity–by-time interaction, with unconstrained covariance between repeated testing time points within the same participant. Contrasts were obtained for the average of the 4-week, 8-week, and 12-week estimates minus baseline. Intent-to-treat methods were followed.

## RESULTS

Baseline characteristics were similar across groups (Table 1). Spatiotemporal outcomes were obtained at 192/220 planned timepoints (HIIT: baseline 26/27, 4-week 25/27, 8-week 23/27, and 12-week 18/27; MAT: baseline 27/28, 4-week 26/28, 8-week 24/28, and 12-week 23/28). Compared with the MAT group, the HIIT group showed significantly greater increases in non-paretic step length (Figures 1-2, Table 2). There were no other significant differences in spatiotemporal changes between the groups.

**Table 1.** Baseline Participant Characteristics. *Values are mean (SD) or N (%)*.

| **Table 1. Baseline Participant Characteristics. Values are mean (SD) or N (%).** |  |  |  |
| --- | --- | --- | --- |
|  | **HIIT**<br>**(N=27)** | **MAT**<br>**(N=28)** | ***p*-value** |
| Age, years | 63.8 (9.9) | 61.5 (9.9) | 0.38 |
| Females, N (%) | 11 (40.7%) | 8 (28.6%) | 0.40 |
| Side of paresis, N (%) |  |  | 1.00 |
| Left | 14 (51.9%) | 14 (50.0%) |  |
| Right | 13 (48.1%) | 14 (50.0%) |  |
| Stroke chronicity, years | 2.7 (1.4) | 2.2 (1.2) | 0.13 |
| Functional ambulation category, 2-4+ | 3.3 (0.6) | 3.4 (0.8) | 0.90 |
| Orthotic device use, N (%) | 12 (44.4%) | 12 (42.9%) | 1.00 |
| Solid ankle foot orthosis | 4 (14.8%) | 2 (7.1%) | 0.42 |
| Articulated/flexible ankle foot orthosis | 7 (25.9%) | 7 (25.0%) | 1.00 |
| Assistive device use, N (%) | 16 (59.3%) | 18 (64.3%) | 0.78 |
| Single point cane | 8 (29.6%) | 11 (39.3%) | 0.33 |
| Narrow-based quad cane | 5 (18.5%) | 1 (3.6%) |  |
| Wide-based quad cane | 3 (11.1%) | 5 (17.9%) |  |
| Front-wheeled walker | 0 (0.0%) | 1 (3.6%) |  |
| Fugl-Meyer lower limb motor score, 0-34 | 23.8 (5.1) | 22.8 (5.1) | 0.44 |
| Self-selected gait speed, m/s | 0.65 (0.29) | 0.62 (0.33) | 0.75 |
| Self-selected gait speed, % predicted | 50.5 (23.3) | 47.3 (25.1) | 0.63 |
| Self-selected gait speed <0.4 m/s, N (%) | 7 (25.9%) | 7 (25.0%) | 1.00 |
| 6-minute walk test, m | 248 (136) | 230 (130) | 0.61 |
| 6-minute walk test, % predicted | 48.5 (26.3) | 44.3 (26.8) | 0.56 |
| *Between-group p-values are from independent t-tests or Fisher exact tests.* |  |  |  |

**Table 2.** Comparison of changes in spatiotemporal gait parameters between short-burst HIIT and MAT in chronic stroke.

|  |  | N |  | Mean HIIT | Mean MAT | Mean Difference<br>(HIIT - MAT) |
| --- | --- | --- | --- | --- | --- | --- |
|  |  | Observations | Observations |  |  |  |
|  |  | HIIT | MAT |  |  |  |
| Non-paretic step length (cm) | Baseline | 26 | 27 | 32.8 [27.5, 38.2] | 32.7 [27.4, 38.0] | 0.1 [-6.9, 7.1] |
| | Mean $\Delta$ | 66 | 73 | 4.4 [1.9, 6.9] | 0.1 [-2.5, 2.7] | 4.3 [1.0, 7.6] |
| Paretic step length (cm) | Baseline | 26 | 27 | 43.5 [38.4, 48.6] | 38.2 [33.1, 43.2] | 5.4 [-1.3, 12.0] |
| | Mean $\Delta$ | 66 | 73 | 4.8 [2.7, 6.8] | 4.6 [2.5, 6.7] | 0.2 [-2.6, 2.9] |
| Step length symmetry | Baseline | 26 | 27 | 76.5 [69.1, 83.8] | 76.2 [68.9, 83.4] | 0.3 [-9.3, 9.9] |
| | Mean $\Delta$ | 66 | 73 | 2.5 [-1.9, 6.9] | -2.5 [-7.0, 2.1] | 5.0 [-0.9, 10.8] |
| Cadence (steps/min) | Baseline | 26 | 27 | 72.4 [67.4, 77.5] | 66.9 [61.9, 71.8] | 5.6 [-1.0, 12.2] |
| | Mean $\Delta$ | 66 | 73 | 9.1 [5.0, 13.2] | 6.4 [2.1, 10.6] | 2.7 [-2.7, 8.2] |
| Paretic Step Ratio | Baseline | 26 | 27 | 59.9 [55.0, 64.8] | 56.6 [51.8, 61.5] | 3.3 [-3.1, 9.7] |
| | Mean $\Delta$ | 66 | 73 | -0.5 [-4.8, 3.8] | 3.7 [-0.7, 8.1] | -4.3 [-9.9, 1.4] |
| Non-paretic step time (s) | Baseline | 26 | 27 | 0.8 [0.7, 0.9] | 0.9 [0.8, 1.0] | -0.1 [-0.3, 0.0] |
| | Mean $\Delta$ | 66 | 73 | -0.2 [-0.3, -0.0] | -0.2 [-0.3, -0.1] | 0.0 [-0.1, 0.2] |
| Paretic step time (s) | Baseline | 26 | 27 | 1.1 [0.9, 1.4] | 1.2 [1.0, 1.4] | -0.1 [-0.3, 0.2] |
| | Mean $\Delta$ | 66 | 73 | -0.1 [-0.2, 0.0] | -0.03 [-0.2, 0.1] | -0.09 [-0.3, 0.1] |
| Step time symmetry | Baseline | 26 | 27 | 79.3 [73.9, 84.8] | 74.3 [68.9, 79.7] | 5.0 [-2.1, 12.2] |
| | Mean $\Delta$ | 66 | 73 | -1.0 [-5.1, 3.2] | 2.8 [-1.5, 7.0] | -3.7 [-9.3, 1.8] |
| Non-paretic single-support time (%) | Baseline | 26 | 26 | 29.8 [28.2, 31.4] | 28.1 [26.5, 29.7] | 1.7 [-0.4, 3.8] |
| | Mean $\Delta$ | 66 | 73 | 4.0 [2.7, 5.4] | 2.5 [1.1, 4.0] | 1.5 [-0.3, 3.3] |
| Paretic single-support time (%) | Baseline | 26 | 26 | 22.0 [20.0, 24.0] | 20.0 [18.0, 22.0] | 2.0 [-0.7, 4.6] |
| | Mean $\Delta$ | 66 | 73 | 1.7 [0.6, 2.8] | 1.6 [0.4, 2.8] | 0.1 [-1.3, 1.6] |
| Single-support symmetry | Baseline | 26 | 26 | 82.2 [78.6, 85.9] | 80.2 [76.6, 83.8] | 2.0 [-2.7, 6.8] |
| | Mean $\Delta$ | 66 | 73 | -1.8 [-4.5, 0.9] | -0.5 [-3.4, 2.4] | -1.3 [-4.9, 2.3] |
| Stride velocity CV (%) | Baseline | 26 | 25 | 10.5 [8.4, 12.7] | 11.7 [9.4, 13.9] | -1.1 [-4.0, 1.7] |
| | Mean $\Delta$ | 66 | 71 | -2.7 [-4.3, -1.1] | -2.6 [-4.3, -0.9] | -0.1 [-2.2, 2.0] |
| Stride time CV (%) | Baseline | 26 | 26 | 8.1 [6.2, 10.0] | 9.6 [7.7, 11.5] | -1.5 [-4.0, 1.0] |
| | Mean $\Delta$ | 66 | 73 | -2.4 [-3.7, -1.0] | -2.3 [-3.7, -0.9] | -0.1 [-1.9, 1.7] |
| Stride length CV (%) | Baseline | 26 | 27 | 7.8 [5.8, 9.8] | 9.2 [7.2, 11.2] | -1.4 [-4.1, 1.2] |
| | Mean $\Delta$ | 65 | 72 | -1.5 [-2.5, -0.5] | -1.4 [-2.5, -0.4] | -0.0 [-1.4, 1.3] |
| Stride velocity SD (cm/s) | Baseline | 26 | 25 | 3.4 [2.8, 4.1] | 3.4 [2.7, 4.1] | 0.0 [-0.9, 0.9] |
| | Mean $\Delta$ | 66 | 71 | 0.2 [-0.5, 0.9] | 0.1 [-0.6, 0.9] | 0.1 [-0.8, 1.1] |
| Stride time SD (s) | Baseline | 26 | 26 | 0.2 [0.1, 0.3] | 0.3 [0.2, 0.3] | -0.1 [-0.2, 0.0] |
| | Mean $\Delta$ | 66 | 73 | -0.07 [-0.12, -0.03] | -0.10 [-0.15, -0.05] | 0.03 [-0.04, 0.09] |
| Stride length SD (cm) | Baseline | 26 | 27 | 4.5 [3.8, 5.2] | 4.8 [4.1, 5.4] | -0.2 [-1.1, 0.6] |
| | Mean $\Delta$ | 65 | 72 | -0.3 [-0.9, 0.3] | -0.3 [-0.9, 0.3] | -0.0 [-0.8, 0.7] |

Abbreviations: HIIT, high-intensity interval training; MAT, moderate intensity aerobic training

**Figure 1.**
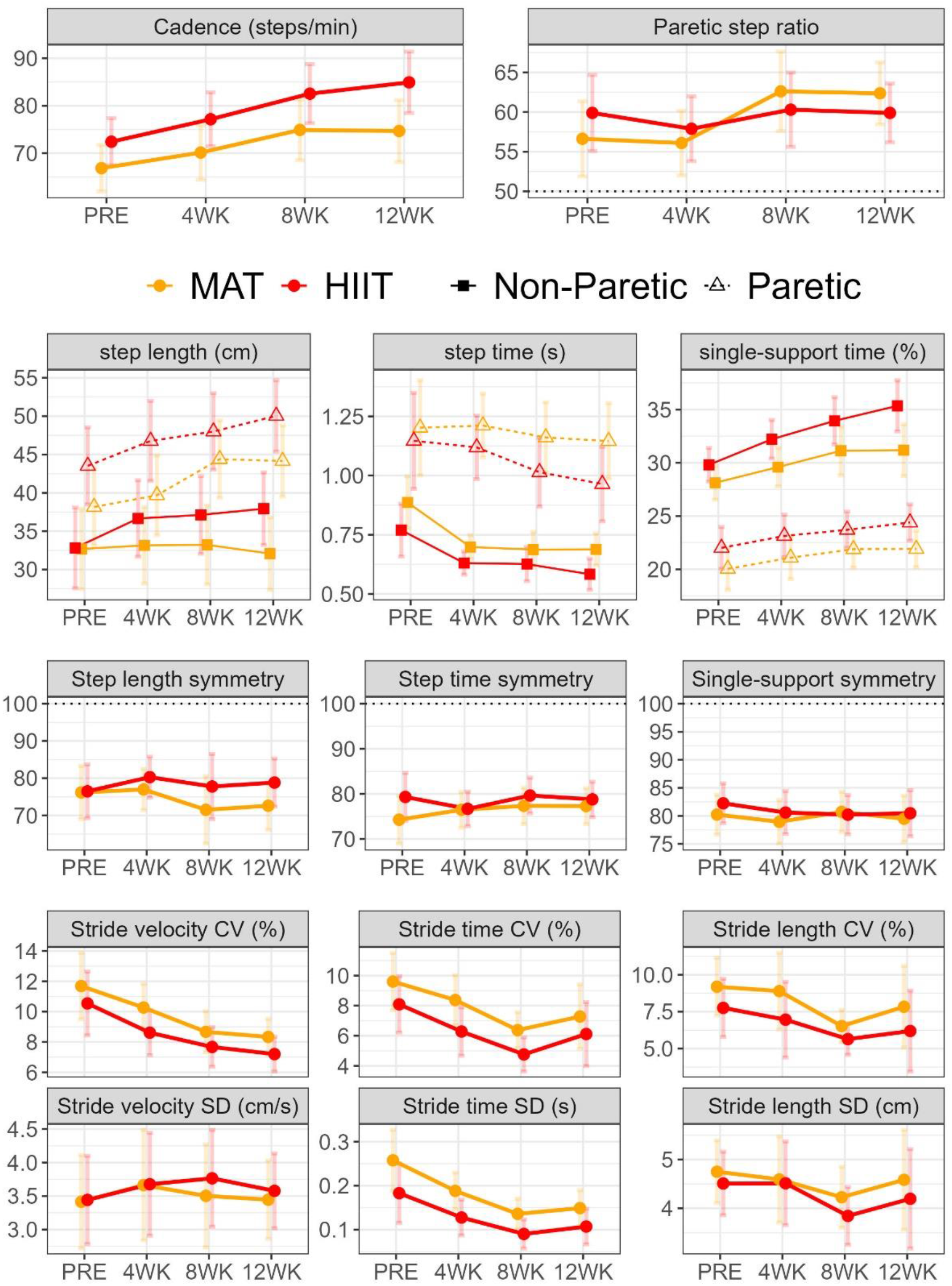
Mean estimates across timepoints (PRE, 4WK, 8WK, 12WK) during 12 weeks of high-intensity interval training (HIIT) or moderate-intensity aerobic training (MAT) in chronic stroke. Error bars are 95% CI. Abbreviations: PRE, pre-testing (baseline); WK, week; CV, coefficient of variation; SD, standard deviation.

**Figure 2.**
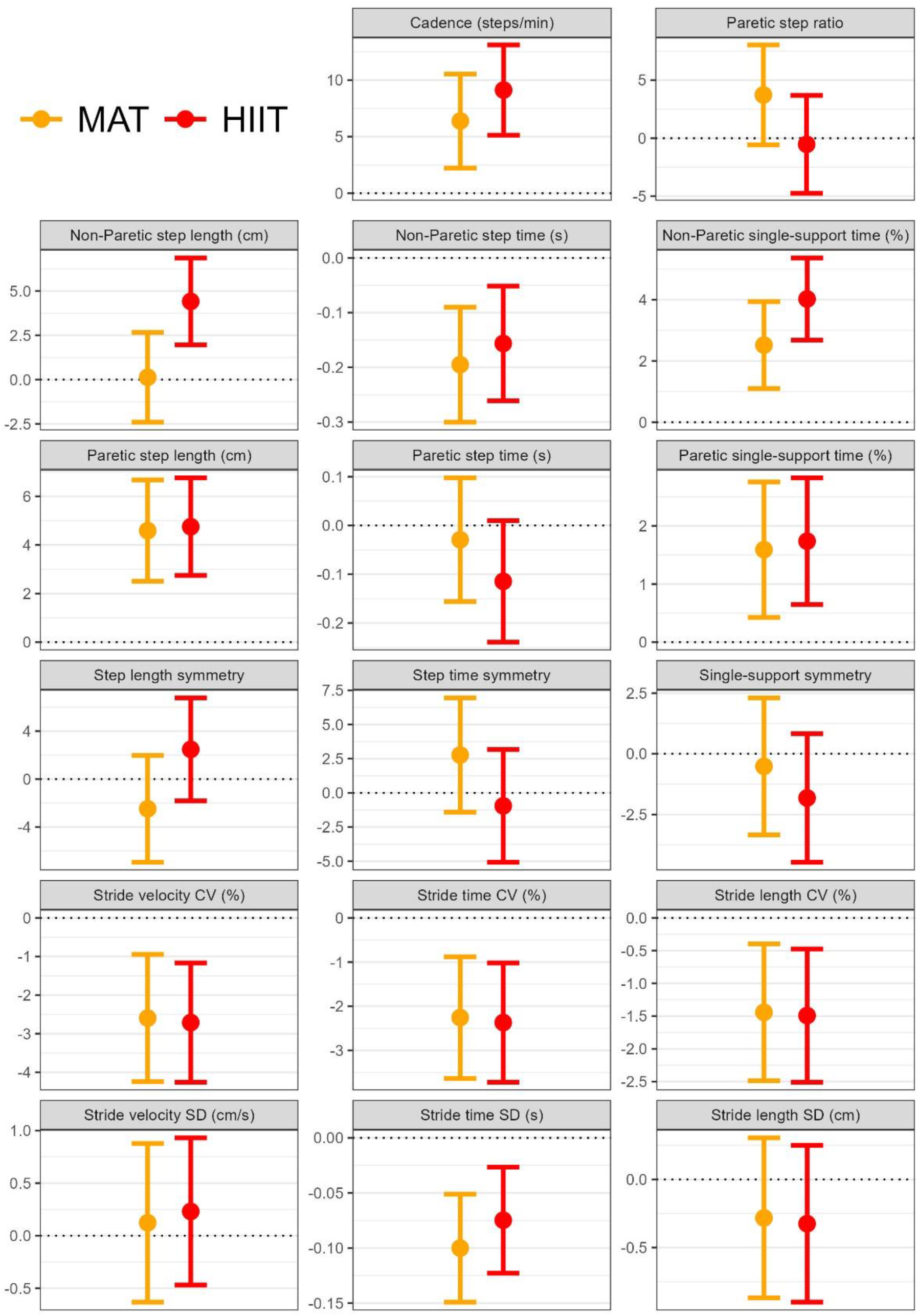
Mean changes from the average of the 4-week, 8-week, and 12-week estimates minus baseline for high-intensity interval training (HIIT) or moderate-intensity aerobic training (MAT) in chronic stroke. Error bars are 95% CI. Abbreviations: CV, coefficient of variation; SD, standard deviation.

Relative to baseline, both groups demonstrated a significant increase in cadence, paretic step length, and bilateral single support time (Figures 1-2, Table 2). Unlike the MAT group, the HIIT group also showed a significant increase in non-paretic step length. Additionally, both groups exhibited significant decreases in the coefficient of variation for stride velocity, stride time, and stride length, and the standard deviation for stride time. No significant changes in spatiotemporal symmetry measures were observed in either group.

## DISCUSSION

In this analysis, we assessed changes in spatiotemporal gait parameters, specifically non-paretic step length, after short-burst HIIT versus MAT in chronic stroke, to compare the effects of maximal versus moderate speed locomotor training. Results indicate maximal speed locomotor training increased non-paretic step length significantly more than MAT, which showed no significant change. Given that non-paretic step length is an indicator of paretic propulsion,^6,10,11^ this finding suggests that short-burst HIIT could increase paretic propulsion more than MAT for the average participant. An increase in paretic propulsion is thought to be an energetically efficient way to increase walking speed, indicating more efficient gait biomechanics and enabling less effortful walking at a given speed.^8,9^ Although future studies are needed for confirmation, this appears to indicate that the faster speeds involved in short-burst HIIT may generally lead to increased biomechanical efficiency. Results also suggest that both moderate and maximal speed locomotor training appear to significantly improve limb-specific spatiotemporal parameters and reduce gait variability in chronic stroke, with no significant impact on spatiotemporal symmetry.

Conversely, we found no significant changes in step length symmetry, single limb support time symmetry, and paretic step ratio in either group (Table 2). However, previous studies have suggested that increased spatiotemporal symmetry may not be an optimal benchmark of biomechanical function, since more symmetrical walking does not necessarily improve metabolic gait efficiency or stability in individuals post-stroke.^9,22-25^ Further, a return to neurotypical gait symmetry is not a universal goal of individuals with stroke,^26-30^ and some “alternative” movement strategies often viewed as “compensation” may actually represent the discovery of adaptive movement solutions given the new constraints placed on the motor control system post-stroke.^31-33^ For example, both HIIT and MAT demonstrated improvements in metabolic gait efficiency^3^ and stability (increased bilateral single-limb support time; Table 2), even in the absence of more symmetrical walking.

In addition to these findings, both groups exhibited significant decreases in relative variability (CV) for stride velocity, stride time, and stride length, and absolute variability (SD) for stride time. Higher gait variability has been correlated with instability and fall risk post-stroke,^34-36^ and people post-stroke commonly demonstrate increased gait variability compared to what is considered an “optimal” level for functional task performance.^33,37^ Therefore, one possible interpretation of the results is that the observed reductions in variability measures may reflect improvements in balance and a reduction in fall risk.^34-36^ However, this inference is based on correlational studies, and it is unclear how much change in variability, if any, may be needed to have a meaningful impact (e.g. on fall risk). Thus, additional research is needed to better interpret the observed changes in gait variability.

It is also important to recognize that motor variability is a task dependent complex construct which is not inherently positive or negative. Reduced variability is often seen as beneficial in stroke recovery as too much variability may indicate neuromuscular instability, poor balance control, and unsteady walking.^34-36,38^ However, too little variability may suggest a lack of flexibility and adaptability in some walking parameters.^39^ People with neurological disability often (counterintuitively) exhibit highly stable motor patterns characterized by deterministic structure (i.e., regularity), as opposed to noisier fluctuations associated with the flexible, adaptive ability to switch more easily among movement patterns.^32,40^ Therefore, while our findings of reduced variability appear to be a positive change in the context of the current study, future research is needed for confirmation.^33,41,42^

## Study Limitations

A primary limitation of this study was the lack of kinetic and kinematic measures, which would have provided a more comprehensive assessment of the underlying biomechanical forces and joint movements associated with the observed improvements in gait. Future studies should include these variables to better understand the differences between HIIT and MAT locomotor training. Another limitation is lack of blinding of personnel who collected the spatiotemporal measures. However, this is less concerning with automated measurements such as these. Additionally, the study was not initially powered for the current ancillary analysis, so it may have been underpowered for some estimates. We also did not control the false discovery rate across the different spatiotemporal measures, since they were each assessing distinct gait features.

## Conclusions

Greater increases in non-paretic step length with short-burst HIIT suggest that training at maximal speeds may yield greater increases in paretic propulsion, a marker of biomechanical efficiency. In addition, both moderate and maximal speed locomotor training (MAT and HIIT) appear to improve other spatiotemporal gait parameters and variability measures in chronic stroke, without significantly altering gait symmetry. Future studies are warranted to explore the impact of maximal speed training on gait mechanics and their relationship to functional outcomes and mobility in individuals with chronic stroke.

## Data Availability

All data produced are available online at https://dash.nichd.nih.gov/study/424597

## Abbreviation List

HIIT: high-intensity interval training
MAT: moderate-intensity aerobic training
HRR: heart rate reserve
CV: coefficient of variation
M-HIT: moderate to high-intensity gait training

## Clinical Trial Registration Number

ClinicalTrials.gov Identifier: NCT03760016

